# Distinct Anaerobic Microbial Signature Differentiates Anal Squamous Cell Carcinoma From High-Grade Squamous Intraepithelial Lesions: A Prospective Metagenomic Study

**DOI:** 10.64898/2026.09.14.26363082

**Authors:** Bryan Nolasco, Travis Lambert, Kai Luo, N. Patrik Brodin, Dean Hosgood, Chandan Guha, Madhur Garg, Shalom Kalnicki, Qibin Qi, Rebecca Levine, Rafi Kabarriti

## Abstract

**Background:** Anal squamous cell carcinoma (ASCC) develops from high-grade squamous intraepithelial lesions (HSIL), but only a subset of patients with HSIL progress to invasive cancer, suggesting that factors beyond persistent human papillomavirus (HPV) infection contribute to disease progression. The anal microbiome may be a contributing factor; however, direct comparisons between HSIL and treatment-naive ASCC remain limited.

**Methods:** Anal canal swabs were collected from adults with histologically confirmed HSIL or nonmetastatic ASCC prospectively enrolled at Montefiore Medical Center from September 2022 through October 2024. Shotgun metagenomic sequencing was performed after human-read removal. Microbial diversity and community composition were compared between disease groups. Differentially abundant species were identified using ANCOM-BC2 and validated using adjusted centered log-ratio regression. Longitudinal changes and MetaCyc pathways associated with persistent ASCC-enriched species were also evaluated. Multivariable analyses adjusted for age, sex, tobacco use, HIV status, and HPV status.

**Results:** Among 121 participants, 102 had HSIL and 19 had ASCC. Observed richness was lower in ASCC (adjusted β=−21.28; p=0.035), while Shannon and inverse Simpson diversity did not differ. Microbial community composition differed by disease group using Bray-Curtis (R²=0.100; p=0.001), Jaccard (R²=0.133; p=0.002), and Aitchison distances (R²=0.172; p=0.001). Of 60 species identified by ANCOM-BC2, 58 were validated, including six enriched in ASCC: *Fusobacterium nucleatum, Parvimonas micra, Peptococcus niger, Porphyromonas asaccharolytica, Porphyromonas levii,* and *Porphyromonas* sp. CAG:1061. All six remained associated with ASCC longitudinally, and five demonstrated greater baseline-to-month-6 decreases in ASCC than HSIL. Twenty-six MetaCyc pathways were linked to the persistent ASCC-enriched species and were also enriched in ASCC, highlighting shared functions in amino acid metabolism, anaerobic degradation, and carbohydrate and energy metabolism.

**Conclusion:** ASCC was characterized by lower observed richness and enrichment of a distinct anaerobic microbial signature that persisted across longitudinal sampling and declined following treatment initiation. These taxa were linked to a shared functional profile dominated by amino acid metabolism and anaerobic metabolic pathways. These findings identify microbial features associated with invasive anal cancer and provide a foundation for evaluating their relationship with disease progression, tumor burden, and treatment response.

## Introduction

Anal squamous cell carcinoma (ASCC) is an uncommon but increasingly prevalent gastrointestinal malignancy, with incidence in the United States rising by approximately 2.7% annually in recent years.^1^ The disease disproportionately affects men who have sex with men (MSM), persons living with HIV (PLWH), and women with a history of genitourinary dysplasia.^2–4^

Similar to cervical cancer, ASCC is primarily driven by persistent infection with high-risk human papillomavirus (HPV), particularly HPV-16 and HPV-18.^5,6^ Progression generally occurs from HPV infection to low-grade and high-grade squamous intraepithelial lesions (HSIL) and ultimately invasive carcinoma. However, most individuals clear HPV infection, and only a subset develop dysplasia or cancer. This indicates that additional factors contribute to anal carcinogenesis.^7^

The microbiome has been associated with cancer development and progression through chronic inflammation, immune dysregulation, DNA damage, and production of biologically active metabolites. ^8–10^ In colorectal cancer, taxa including *Fusobacterium nucleatum, enterotoxigenic Bacteroides fragilis,* and *pks-positive Escherichia coli* have been linked to proinflammatory signaling, genotoxin production, and suppression of antitumor immune responses. ^8,11^ Similar associations have been reported in HPV-related cervical disease, where depletion of protective Lactobacillus species and enrichment of anaerobic taxa such as Gardnerella, Prevotella, and Sneathia have been associated with HPV persistence and progression to high-grade dysplasia and invasive cancer.^12,13^ These findings support further evaluation of the microbiome in anal carcinogenesis.

Several studies have identified microbiome differences across the spectrum of anal disease. Elnaggar et al. (2023) compared anorectal samples from high-risk individuals and patients with anal dysplasia or cancer and found enrichment of *Peptoniphilus, Fusobacterium, Porphyromonas,* and *Prevotella* in cancer samples.^14^ In HIV-positive men who have sex with men, Brickman et al. (2024) found that HSIL was associated with enrichment of proinflammatory and potentially carcinogenic taxa and depletion of short-chain fatty acid-producing commensals compared with lesion-free controls.^15^ More recently, tissue-based metatranscriptomic analyses identified increased abundance and activity of *Fusobacterium nucleatum* and *Bacteroides fragilis* in ASCC compared with precancerous lesions, together with bacterial enzymes and metabolites that may contribute to carcinogenesis.^16^

Despite these advances, important gaps remain. Most prior studies compared dysplasia or cancer with high-risk or normal controls, whereas direct comparisons between HSIL and ASCC remain limited. Many studies have also included relatively small cohorts, limiting the ability to identify reproducible differences across disease states. We therefore prospectively compared the anal microbiome of patients with HSIL and non-metastatic ASCC and evaluated longitudinal changes in the identified microbial signature during and after treatment.

## Methods

### Study Design and Setting

This study was conducted as a prespecified microbiome analysis within an ongoing prospective clinical protocol evaluating HPV-associated anal disease at Montefiore Medical Center in the Bronx, New York. The present study consisted of a baseline comparison of anal microbiome composition between patients with HSIL and newly diagnosed, non-metastatic, treatment-naïve ASCC, followed by longitudinal evaluation of microbiome changes during and after treatment. Anal canal swab specimens included in this analysis were collected at the time of participant consent between September 2022 and October 2024. All microbiome-related objectives, specimen collection procedures, and analytic plans were defined a priori within the approved clinical protocol and conducted in accordance with institutional review board approval. Written informed consent was obtained from all participants prior to specimen collection.

### Study Population and Cohort Definitions

Participants were identified and enrolled at the Montefiore Medical Center. Eligible patients were included if they were ≥ 18 years of age, had an available baseline anal canal swab specimen collected during the defined study period, and had histologically confirmed HSIL or ASCC. Participants were excluded if they had received prior chemoradiation or surgical treatment for anal cancer before baseline sampling or had evidence of metastatic anal cancer at diagnosis. Samples with inadequate sequencing depth, failed sequencing quality control procedures, or unavailable sequencing data were excluded from downstream analyses.

Baseline demographic and clinical variables, including age, sex, race, ethnicity, HIV status, and tobacco use, were obtained from routine clinical documentation and electronic medical record review at the time of enrollmentTobacco use was categorized as never versus ever use based on self-reported social history. HIV status was documented from the medical record and treated as a binary variable (positive vs negative). Anal HPV status was determined using anal swab testing and classified as HPV positive or negative. Detailed HPV genotyping included HPV 16, HPV 18, HPV 45, and other high-risk HPV strains, when available from anal cytology testing performed as part of routine clinical care. Disease stage was applicable only to the ASCC cohort and was determined at diagnosis using AJCC staging guidelines.^17^

Participants were classified into two analytic cohorts based on histopathology and clinical status at baseline. Baseline was defined as the timepoint at which the anal canal swab specimen was collected for microbiome analysis. For HSIL participants, baseline corresponded to the enrollment visit. For ASCC participants, baseline corresponded to the pretreatment visit prior to initiation of chemoradiation, ensuring that microbiome profiles reflected untreated disease.

### Specimen Collection and Sequencing

Anal canal specimens were collected at enrollment using a sterile dual polyurethane foam swab system (BD BBL CultureSwab EZ). During anal examination, the swab was inserted into the anal canal and rotated approximately 5–10 times along the anal canal wall to obtain the specimen, without tissue removal or biopsy. Swabs were immediately placed into sterile collection tubes, transported on ice, and stored at −20 °C until processing. DNA extraction and shotgun metagenomic sequencing were performed by CosmosID using validated laboratory pipelines. Total microbial DNA was extracted from anal swab specimens using bead beating and chemical lysis followed by extraction with the QIAGEN DNeasy PowerSoil Pro Kit. DNA quantity and quality were assessed prior to library preparation. Sequencing libraries were prepared using the Watchmaker DNA Library Prep Kit with IDT xGen UDI primers and IDT Stubby Adapters, followed by paired-end sequencing (2 × 150 bp) on the Illumina NovaSeq X platform. Positive and negative controls were included during sequencing and quality control procedures to monitor for contamination and sequencing performance. Raw sequencing data were returned to the study team for downstream bioinformatic processing.

### Sequence Processing and Taxonomic Profiling

Raw sequencing reads underwent preprocessing, quality control, and host read removal using KneadData with default parameters.^18^ Low-quality bases, sequencing adapters, and low-complexity reads were removed prior to downstream analysis, and human DNA was depleted by alignment to the human reference database. ^19^ Only non-host microbial reads were retained for downstream analyses. Summary metrics of sequencing depth, read retention, and alignment rates were reviewed to ensure adequate sequencing quality across samples. Microbial taxonomic profiling was performed using the Woltka pipeline,^20^ which maps shotgun metagenomic reads to the Web of Life (WoL) reference database^21^ using Bowtie2 alignment.^22^ Reads were aligned using Bowtie2 and classified into taxonomic units using a tree-structured reference framework. Species-level feature tables were generated and imported into R as phyloseq objects for downstream analysis.^23^ In the main analyses, we focused on core bacterial species with a minimum relative abundance of 0.01% in at least 20% of all baseline HSIL and ASCC samples, resulting in 447 species for downstream analysis.

### Statistical Analysis

Baseline demographic and clinical characteristics were summarized separately for participants with HSIL and ASCC. Continuous variables were summarized using means with standard deviations, and categorical variables were summarized using frequencies and percentages. Between-group comparisons were performed using parametric or nonparametric tests for continuous variables and chi-square or Fisher’s exact tests for categorical variables, as appropriate. Unless otherwise specified, all statistical tests were two-sided, and statistical significance was defined as a *p* value < 0.05.

Analyses were conducted in R version 4.5.2 using publicly available packages, including phyloseq, microbiome, vegan, ANCOMBC, microbiomeMarker, and caret. Fixed seed values were set before model training to support reproducibility. Because of the limited sample size, leave-one-out cross-validation was used for random forest analyses.

### Baseline analyses

We initially examined differences in microbial diversity between disease status. For alpha diversity, observed species richness, Shannon diversity, and inverse Simpson diversity were calculated from raw count data. For beta diversity, Bray–Curtis dissimilarity, Jaccard distance, and Aitchison distance were calculated and further visualized using principal coordinates analysis. The relationship between disease status and the overall microbial composition was calculated via PERMANOVA (999 permutations) adjusting for age, sex, tobacco use, HIV status, and HPV status.

Species-level differential abundance between HSIL and ASCC at baseline was evaluated using Analysis of Compositions of Microbiomes with Bias Correction 2 (ANCOM-BC2) applied to baseline core-filtered count data. Models were adjusted for age, sex, tobacco use, HIV status, and HPV status. Species identified as significant by ANCOM-BC2 were subsequently evaluated using multivariable linear regression models of centered log-ratio-transformed abundance. These models were adjusted for age, sex, tobacco use, HIV status, and HPV status. Positive coefficients indicated enrichment in ASCC, whereas negative coefficients indicated enrichment in HSIL. Final validated candidate species were defined as taxa that were statistically significant using both ANCOM-BC2 and CLR regression and demonstrated consistent directionality across the two approaches. Multiple comparisons were controlled using the false discovery rate (FDR).

Linear discriminant analysis effect size was used as an exploratory complementary method to identify species associated with disease group. Random forest classifiers were also trained using species-level abundance profiles to evaluate the ability of baseline microbial features to distinguish ASCC from HSIL. Model performance was evaluated using leave-one-out cross-validation, with discrimination summarized by the area under the receiver operating characteristic curve and its 95% confidence interval. Feature-importance rankings were extracted to identify taxa contributing most strongly to classification. Given the modest sample size and potential risk of overfitting, LEfSe and random forest analyses were considered hypothesis-generating and were not used for primary candidate-species selection.

### Longitudinal analyses

Fifty-eight validated baseline candidate species were included in the longitudinal dataset. Repeated measurements obtained at baseline, Month 6, and Month 12 were analyzed using linear mixed-effects models with participant-specific random intercepts to account for within-participant correlation. Month 2 samples were excluded from the primary between-group longitudinal models because they were available only for participants with ASCC. Models evaluated whether CLR-transformed abundances of the validated baseline species remained different between HSIL and ASCC across repeated sampling after adjustment for age, sex, tobacco use, HIV status, and HPV status. Regression coefficients, 95% confidence intervals, and *p* values were reported.

Changes from baseline to Month 6 were evaluated using delta-CLR values, calculated as the Month 6 CLR-transformed abundance minus the corresponding baseline value for each participant. Between-group differences in temporal change were evaluated for the validated baseline candidate species. Negative regression coefficients indicated a greater decrease over time in ASCC relative to HSIL.

Because Month 2 specimens were available only for participants with ASCC, paired analyses were performed among 14 ASCC participants to evaluate baseline-to-Month 2 changes across the 58 validated baseline species using unadjusted change-score linear models. Regression coefficients, 95% confidence intervals, and p values were reported, with negative coefficients indicating decreased CLR-transformed abundance from baseline to Month 2.

### Community-Level Functional Pathway Analysis

To characterize community-level functional patterns associated with the persistent ASCC-enriched microbial signature, MetaCyc pathway abundances were evaluated in relation to the species that remained enriched in ASCC across longitudinal sampling. Pathways detected in at least 20% of samples were CLR transformed. For each species, multivariable linear regression models evaluated associations between species and pathway CLR abundances after adjustment for age, sex, tobacco use, HIV status, and HPV status. Separate adjusted models evaluated pathway enrichment in ASCC versus HSIL. False discovery rates were controlled using the Benjamini-Hochberg method. Primary species-pathway associations were defined as positive associations in which the pathway was also enriched in ASCC (FDR-adjusted q < 0.05 in both models). Because pathway abundances were measured at the community level, these analyses did not demonstrate species-specific functional contributions or causality.

## Results

### Cohort and Baseline Characteristics

A total of 121 participants with evaluable baseline anal canal microbiome samples were included in the primary baseline analyses, comprising 102 patients with HSIL and 19 patients with ASCC (Table 1). One additional HSIL participant and three additional ASCC participants without usable baseline samples contributed follow-up specimens and were included only in longitudinal analyses, yielding a total longitudinal cohort of 125 participants. Patients with ASCC were older than those with HSIL (62.4 vs 52.8 years, p = 0.023). Baseline demographic characteristics were otherwise generally comparable between groups. HIV positivity was higher among HSIL patients than ASCC patients (90.2% vs 52.6%, p < 0.001). HPV positivity was high in both groups (90.2% vs 84.2%, p = 0.428) (Table 1).

**Table 1.** Baseline Demographic and Clinical Characteristics of Patients.

| Variable | Total<br>(n = 121) | ASCC<br>(n = 19) | HSIL<br>(n = 102) | p-value |
| --- | --- | --- | --- | --- |
| **Age, years, mean (SD)** | 54.3 (12.6) | 62.4 (16.4) | 52.8 (11.3) | **0.023** |
| **Sex** |  |  |  | 0.239 |
| Male | 69 (57.0%) | 8 (42.1%) | 61 (59.8%) |  |
| Female | 52 (43.0%) | 11 (57.9%) | 41 (40.2%) |  |
| **Race** |  |  |  | 0.743 |
| Black or African American | 51 (42.1%) | 7 (36.8%) | 44 (43.1%) |  |
| White | 10 (8.3%) | 1 (5.3%) | 9 (8.8%) |  |
| Asian | 1 (0.8%) | 0 (0%) | 1 (1.0%) |  |
| Other | 47 (38.8%) | 10 (52.6%) | 37 (36.3%) |  |
| Declined or Unknown | 12 (9.9%) | 1 (5.3%) | 11 (10.8%) |  |
| **Ethnicity** |  |  |  | 0.771 |
| Hispanic/Latino | 51 (42.1%) | 9 (47.4%) | 42 (41.2%) |  |
| Not Hispanic/Latino | 57 (47.1%) | 9 (47.4%) | 48 (47.1%) |  |
| Declined or Unknown | 13 (10.7%) | 1 (5.3%) | 12 (11.8%) |  |
| **Tobacco Use** |  |  |  | 0.316 |
| Never | 48 (39.7%) | 10 (52.6%) | 38 (37.3%) |  |
| Ever-use | 73 (60.3%) | 9 (47.4%) | 64 (62.7%) |  |
| **HIV Status** |  |  |  | **<0.001** |
| Negative | 19 (15.7%) | 9 (47.4%) | 10 (9.8%) |  |
| Positive | 102 (84.3%) | 10 (52.6%) | 92 (90.2%) |  |
| **HPV Status** |  |  |  | **0.428** |
| HPV negative | 13 (10.7%) | 3 (15.8%) | 10 (9.8%) |  |
| HPV positive | 108 (89.3%) | 16 (84.2%) | 92 (90.2%) |  |
| **HPV Genotype (HPV+ only)** |  |  |  |  |
| HPV 16 and/or 18 | 54 (50.0%) | 14 (87.5%) | 40 (43.5%) |  |
| Other HPV types | 54 (50.0%) | 2 (12.5%) | 52 (56.5%) |  |
| **Disease Stage** |  |  |  |  |
| Stage 1 |  | 2 (10.5%) |  |  |
| Stage 2 |  | 7 (36.8%) |  |  |
| Stage 3 |  | 10 (52.6%) |  |  |

*Baseline characteristics of participants with HSIL and ASCC included in the primary baseline analysis (n = 121; HSIL, n = 102; ASCC, n = 19). Continuous variables were compared using Welch's two-sample t test, and categorical variables using chi-square or Fisher's exact tests, as appropriate. HPV genotype percentages were calculated among HPV-positive participants only. Abbreviations: ASCC, anal squamous cell carcinoma; HIV, human immunodeficiency virus; HPV, human papillomavirus; HSIL, high-grade squamous intraepithelial lesion.*

### Baseline Alpha Diversity and Community Structure

Alpha diversity was assessed using observed richness, Shannon diversity, and inverse Simpson diversity at the species level. Observed richness was lower in ASCC compared with HSIL in unadjusted analyses (Wilcoxon p = 0.0083), while Shannon diversity was similar between groups (Wilcoxon p = 0.78) (Figure 1). This pattern persisted after covariate adjustment, with ASCC associated with lower observed richness (β = −21.28, 95% CI −41.01 to −1.55, p = 0.035), while Shannon diversity (β = 0.01, 95% CI −0.38 to 0.40, p = 0.949) and inverse Simpson diversity (β = 0.90, 95% CI −5.70 to 7.49, p = 0.789) did not differ significantly between groups (Figure 1; Supplementary Figure 1).

**Figure 1.**
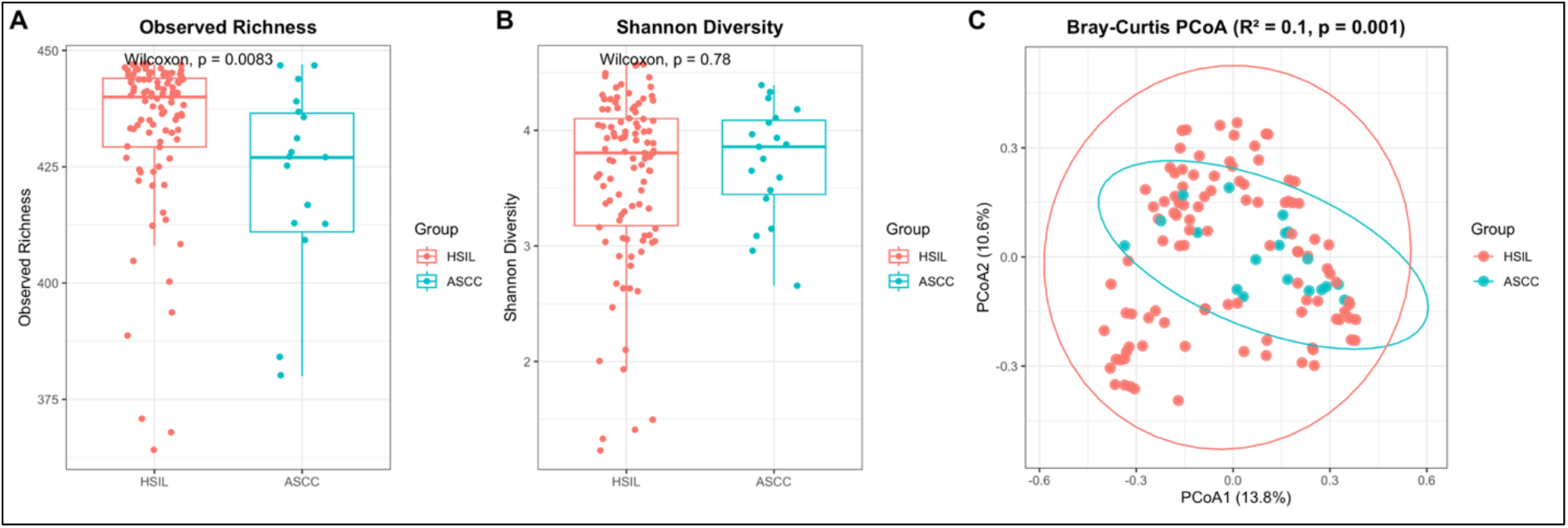
Alpha diversity and microbial community composition differ between HSIL and ASCC. (A) Observed species richness and (B) Shannon diversity in baseline anal canal microbiome samples from patients with high-grade squamous intraepithelial lesions (HSIL; n = 102) and anal squamous cell carcinoma (ASCC; n = 19). Boxplots show the median and interquartile range, with individual samples overlaid. Between-group comparisons were performed using Wilcoxon rank-sum tests. Observed richness was significantly lower in ASCC, whereas Shannon diversity did not differ between groups. (C) Principal coordinates analysis (PCoA) of Bray-Curtis dissimilarity demonstrates differences in overall microbial community composition between HSIL and ASCC. Each point represents an individual baseline sample and ellipses depict the distribution of samples within each disease group. Disease-associated differences in community composition were evaluated using PERMANOVA after adjustment for age, sex, tobacco use, HIV status, and HPV status (R² = 0.100, p = 0.001).

Overall microbial community composition differed between HSIL and ASCC across multiple beta diversity metrics. In adjusted PERMANOVA models, Bray–Curtis dissimilarity demonstrated significant separation by disease group (R² = 0.100, p = 0.001) (Figure 1). Similar findings were observed using Jaccard distance (R² = 0.133, p = 0.002) and Aitchison distance derived from CLR-transformed abundances (R² = 0.172, p = 0.001) (Supplementary Figure 2). PERMDISP testing demonstrated no significant dispersion differences for Bray–Curtis (p = 0.971) or Aitchison distances (p = 0.925), supporting that these findings reflected differences in community structure rather than dispersion alone. Jaccard dispersion differed between groups (p = 0.015) and was therefore interpreted more cautiously (Supplementary Figure 3). Covariate-adjusted constrained ordination using Bray–Curtis distances further demonstrated disease-associated community separation (p = 0.001) (Supplementary Figure 4).

### Baseline Differential Abundance Discovery

Species-level differential abundance analyses identified a broad set of taxa that differed between HSIL and ASCC after covariate adjustment. ANCOM-BC2 identified 60 FDR-significant species, including 7 ASCC-enriched and 53 HSIL-enriched taxa (Figure 2; Supplementary Table 2). ASCC-enriched taxa were dominated by an anaerobic cluster that included *Fusobacterium nucleatum*, *Parvimonas micra*, *Porphyromonas levii*, *Porphyromonas asaccharolytica*, *Porphyromonas* sp. CAG:1061, *Peptococcus niger*, and *Acidaminococcus intestini* CAG:325 (Figure 2; Supplementary Table 2). In contrast, microbial features of HSIL were characterized by enrichment of multiple *Prevotella* species and enteric-associated taxa, including *Escherichia coli*, *Shigella flexneri*, *Shigella dysenteriae*, *Salmonella enterica*, *Citrobacter amalonaticus*, *Citrobacter freundii*, *Enterobacter cloacae*, and several *Kluyvera* species (Figure 2; Supplementary Table 2).

**Figure 2.**
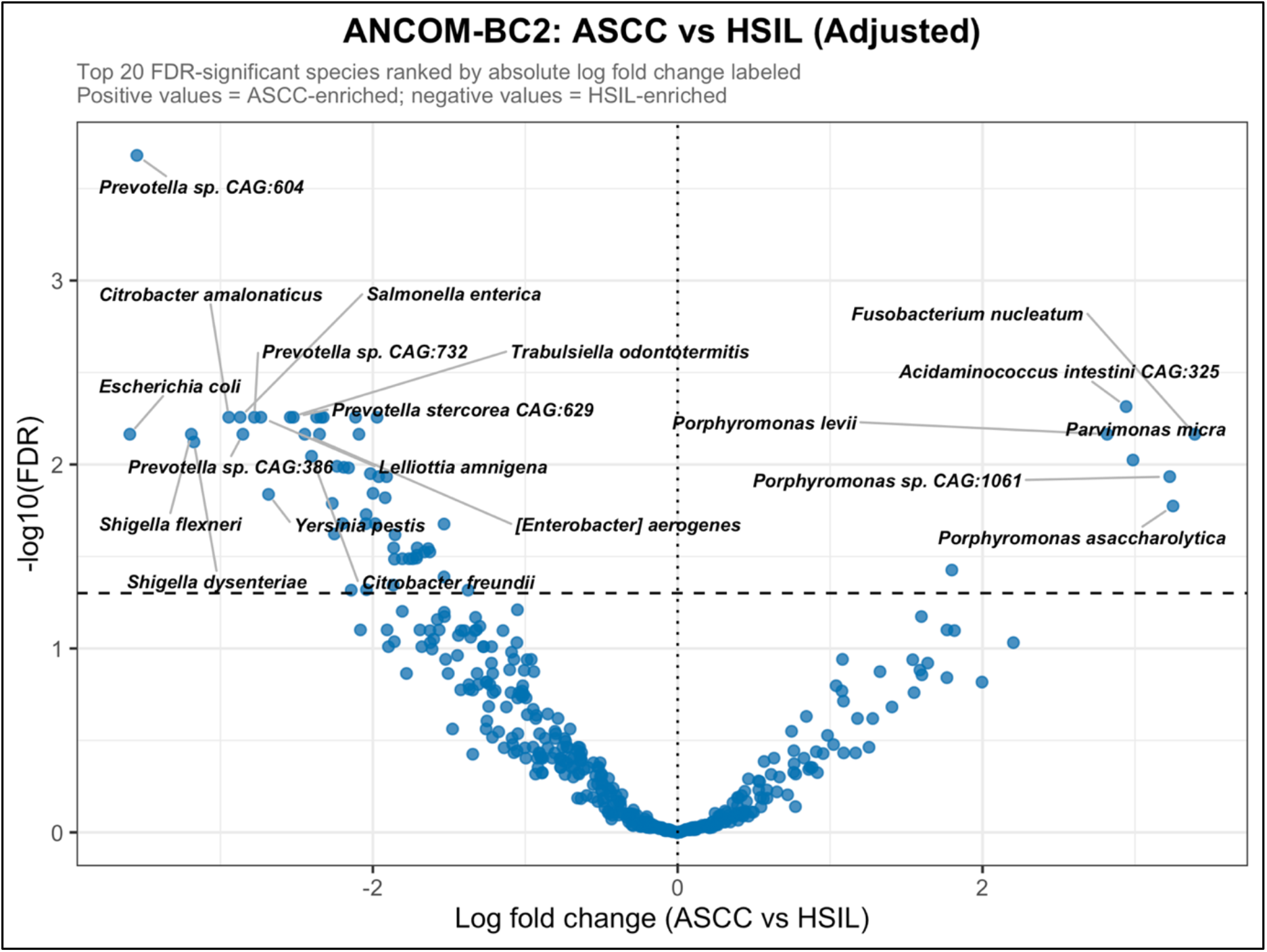
Covariate-adjusted differential abundance analysis identifies distinct microbial species associated with ASCC and HSIL. Volcano plot showing species-level differential abundance between ASCC and HSIL using ANCOM-BC2. Models were adjusted for age, sex, tobacco use, HIV status, and HPV status. The x-axis represents log fold change for ASCC relative to HSIL, with positive values indicating enrichment in ASCC and negative values indicating enrichment in HSIL. The y-axis represents −log10 of the FDR-adjusted q value. The 20 FDR-significant species with the largest absolute log fold changes are labeled. The horizontal dashed line represents an FDR-adjusted q value of 0.05, and the vertical dashed line indicates no difference between groups. ANCOM-BC2 identified 60 FDR-significant species, including 7 enriched in ASCC and 53 enriched in HSIL.

### Validation of Candidate Species Using CLR Regression

To refine the baseline microbial signature, ANCOM-BC2 significant taxa were tested in covariate-adjusted CLR regression models. Of the 60 ANCOM-BC2 significant species, 58 remained significant after CLR validation with consistent directionality across models, including 6 ASCC-enriched and 52 HSIL-enriched taxa (Figure 3; Supplementary Table 3). Species associated with ASCC showing consistent associations in these two models included *Fusobacterium nucleatum* (β = 4.32, 95% CI 2.36 to 6.27, q < 0.001), *Porphyromonas* sp. CAG:1061 (β = 3.74, 95% CI 2.10 to 5.38, q < 0.001), *Porphyromonas asaccharolytica* (β = 3.53, 95% CI 1.88 to 5.19, q < 0.001), *Porphyromonas levii* (β = 3.41, 95% CI 2.16 to 4.66, q < 0.001), *Parvimonas micra* (β = 3.37, 95% CI 1.93 to 4.82, q < 0.001), and *Peptococcus niger* (β = 1.67, 95% CI 0.37 to 2.96, q = 0.0174) (Figure 3). Validated HSIL-enriched taxa included *Salmonella enterica* (β = −2.73, 95% CI −4.18 to −1.28, q = 0.0028), *Citrobacter amalonaticus* (β = −2.63, 95% CI −4.10 to −1.15, q = 0.0033), *Escherichia coli* (β = −3.07, 95% CI −4.90 to −1.25, q = 0.0041), *Shigella flexneri* (β = −3.06, 95% CI −4.89 to −1.23, q = 0.0041), and *Shigella dysenteriae* (β = −3.03, 95% CI −4.85 to −1.20, q = 0.0042) (Figure 3; Supplementary Table 3). Only two ANCOM-BC2 significant taxa did not validate in CLR regression: *Allisonella histaminiformans* and *Acidaminococcus intestini* CAG:325. Meanwhile, the majority of these identified species were recaptured by significant results in LEfSe (Supplementary Figure 5). A random forest classifier demonstrated good discriminatory performance for distinguishing HSIL from ASCC (AUC = 0.844, 95% CI 0.750 to 0.938). Highly ranked features included *Porphyromonas somerae*, *Porphyromonas asaccharolytica*, *Campylobacter ureolyticus*, *Porphyromonas* sp. CAG:1061, *Porphyromonas levii*, and *Parvimonas micra*, several of which overlapped with the validated ANCOM-BC2/CLR signature (Supplementary Figure 6).

**Figure 3.**
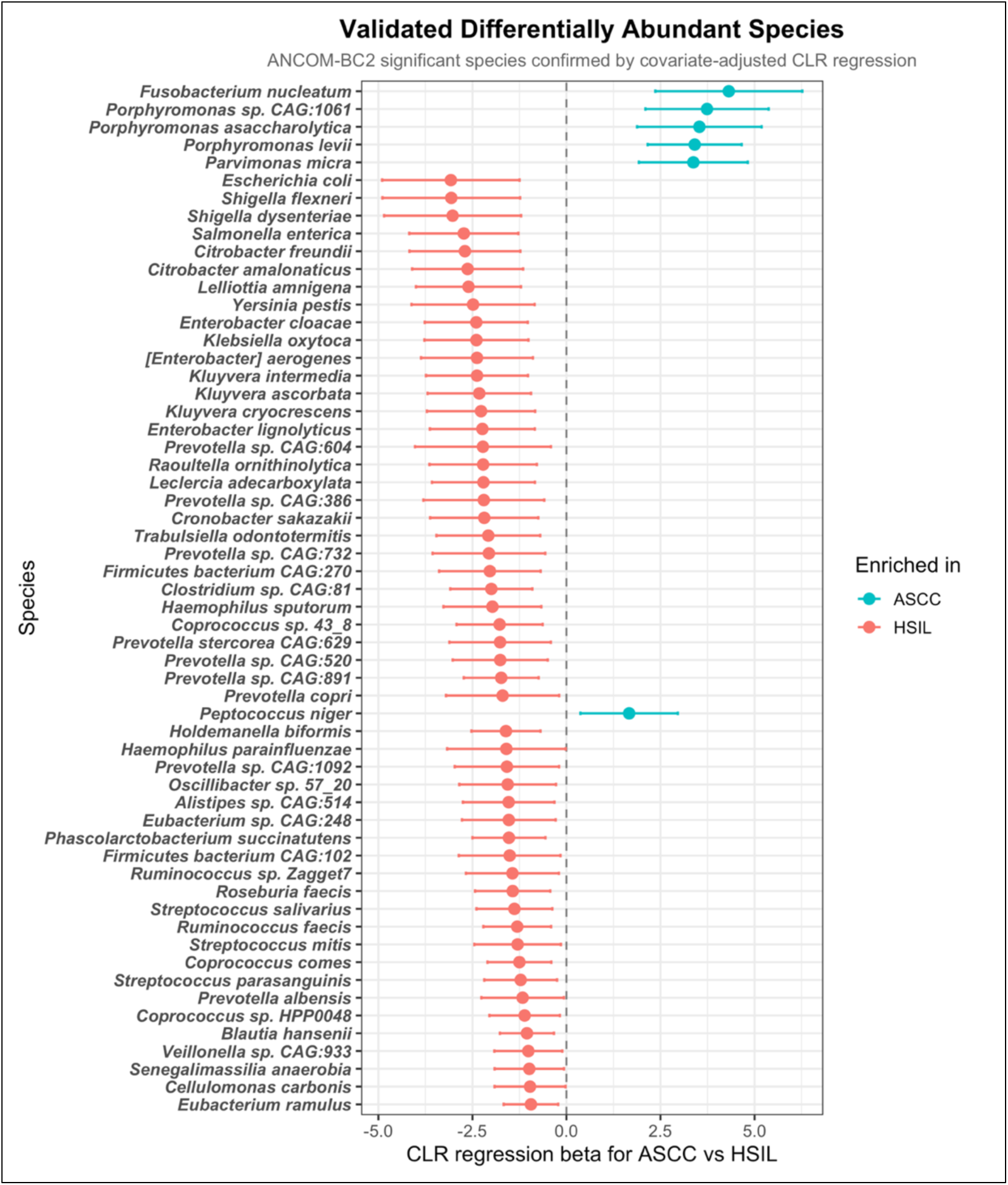
Covariate-adjusted CLR regression validates the baseline disease-associated microbial signature. Forest plot showing centered log-ratio (CLR) regression coefficients and 95% confidence intervals for species initially identified as significant by ANCOM-BC2 and subsequently evaluated using covariate-adjusted CLR regression. Models were adjusted for age, sex, tobacco use, HIV status, and HPV status. Of 60 species identified by ANCOM-BC2, 58 remained significant with consistent directionality across both approaches. Positive coefficients indicate enrichment in ASCC, whereas negative coefficients indicate enrichment in HSIL. Points represent regression coefficients and horizontal lines represent 95% confidence intervals. Six validated species were enriched in ASCC: *Fusobacterium nucleatum, Porphyromonas* sp. CAG:1061, *Porphyromonas asaccharolytica, Porphyromonas levii, Parvimonas micra,* and *Peptococcus niger*. The vertical dashed line at β = 0 indicates no difference between groups.

### Longitudinal Persistence of Validated Baseline Species

Longitudinal analyses were then restricted to the 58 validated baseline candidate species to determine whether baseline disease-associated taxa remained differentially abundant over repeated sampling. Across Baseline, Month 6, and Month 12 samples, 11 species remained significantly associated with disease group in adjusted linear mixed-effects models, including 6 ASCC-enriched and 5 HSIL-enriched taxa (Figure 4; Supplementary Table 4). The persistent ASCC-enriched taxa were the same core anaerobic species identified at baseline: *Fusobacterium nucleatum* (β = 3.76, 95% CI 2.11 to 5.41, p < 0.001), *Porphyromonas* sp. CAG:1061 (β = 3.28, 95% CI 1.87 to 4.70, p < 0.001), *Porphyromonas levii* (β = 3.12, 95% CI 1.97 to 4.26, p < 0.001), *Parvimonas micra* (β = 2.94, 95% CI 1.76 to 4.12, p < 0.001), *Porphyromonas asaccharolytica* (β = 2.58, 95% CI 1.13 to 4.03, p < 0.001), and *Peptococcus niger* (β = 2.13, 95% CI 0.96 to 3.30, p < 0.001) (Figure 4). The persistent HSIL-enriched taxa were *Escherichia coli* (β = −1.49, 95% CI −2.68 to −0.30, p = 0.0142), *Shigella flexneri* (β = −1.48, 95% CI −2.67 to −0.28, p = 0.0157), *Shigella dysenteriae* (β = −1.46, 95% CI −2.65 to −0.28, p = 0.0162), *Salmonella enterica* (β = −1.00, 95% CI −1.88 to −0.12, p = 0.0269), and *Citrobacter amalonaticus* (β = −0.92, 95% CI −1.82 to −0.02, p = 0.0449) (Figure 4). Overall, these findings show that the most reproducible longitudinal signal was concentrated in the same ASCC-enriched anaerobic cluster identified at baseline, while many baseline HSIL-associated enteric taxa attenuated over repeated sampling.

**Figure 4.**
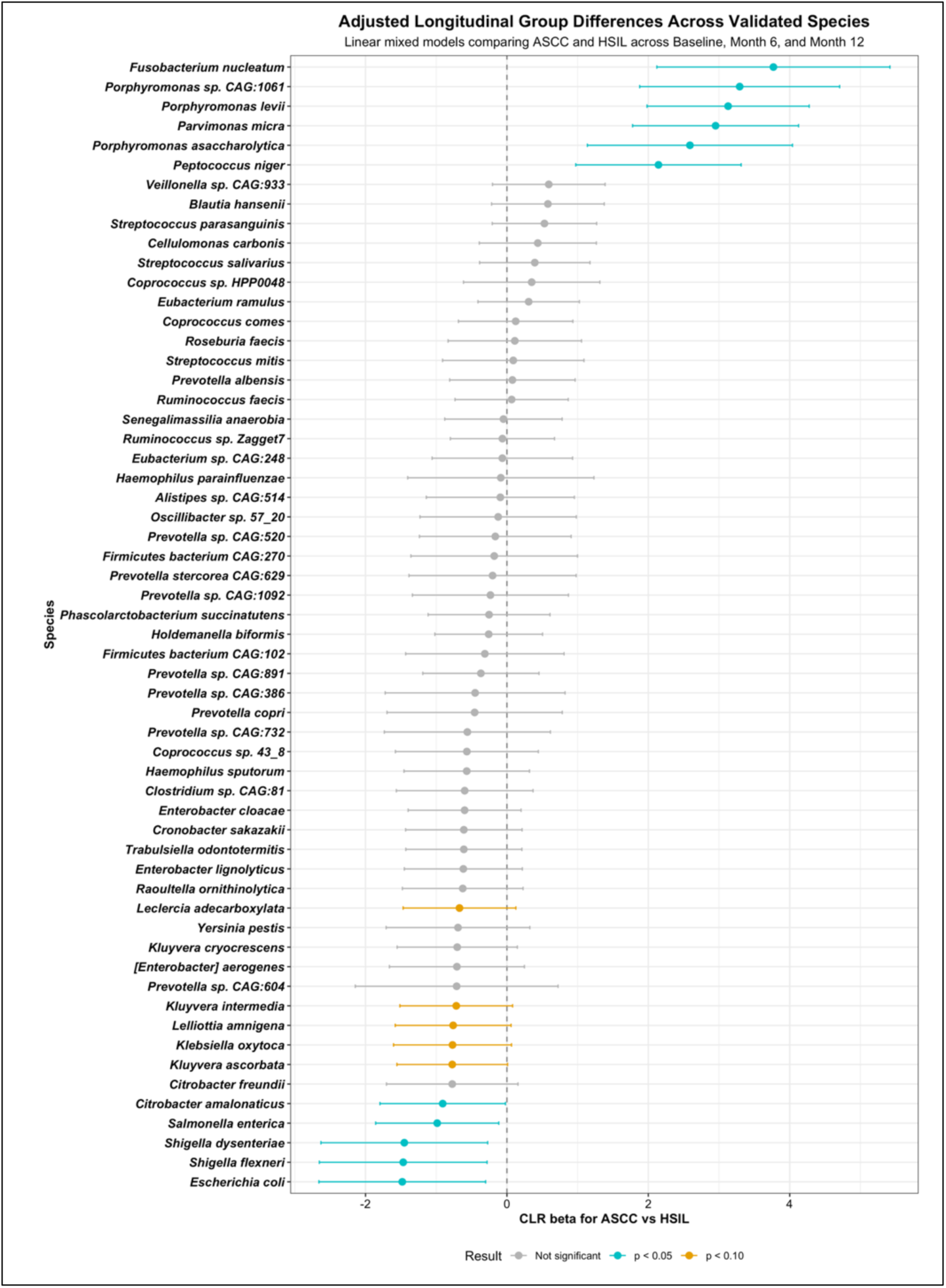
Longitudinal persistence of validated disease-associated microbial species. Forest plot showing adjusted differences in centered log-ratio (CLR) abundance between ASCC and HSIL across baseline, Month 6, and Month 12. The 58 validated baseline candidate species were evaluated using linear mixed-effects models with participant-specific random intercepts to account for repeated measurements. Models were adjusted for age, sex, tobacco use, HIV status, and HPV status. Points represent regression coefficients for ASCC relative to HSIL and horizontal lines represent 95% confidence intervals. Positive coefficients indicate greater abundance in ASCC, whereas negative coefficients indicate greater abundance in HSIL. Eleven species remained significantly associated with disease group, including all six validated ASCC-enriched anaerobic species and five HSIL-enriched species. Colors indicate associations with p < 0.05, p < 0.10, or nonsignificant associations, as indicated in the figure. The vertical dashed line at β = 0 indicates no difference between disease groups.

### Longitudinal Change from Baseline to Month 6

Because the strongest ASCC-associated species remained elevated longitudinally, we next evaluated whether their abundance changed differentially over time. In delta CLR analyses comparing Baseline-to-Month 6 change between groups, five validated ASCC-enriched species demonstrated significantly greater decreases in ASCC relative to HSIL: *Fusobacterium nucleatum* (β = −4.01, 95% CI −6.74 to −1.28, p = 0.0046), *Porphyromonas asaccharolytica* (β = −3.60, 95% CI −5.58 to −1.63, p < 0.001), *Parvimonas micra* (β = −2.72, 95% CI −4.81 to −0.63, p = 0.0114), *Porphyromonas* sp. CAG:1061 (β = −2.33, 95% CI −4.34 to −0.31, p = 0.0243), and *Porphyromonas levii* (β = −2.10, 95% CI −3.93 to −0.27, p = 0.0251) (Figure 5). Thus, the same anaerobic taxa that were most strongly enriched in untreated ASCC at baseline showed the clearest decreases after treatment initiation, supporting a tumor-associated microbial signature that partially attenuates over time.

**Figure 5.**
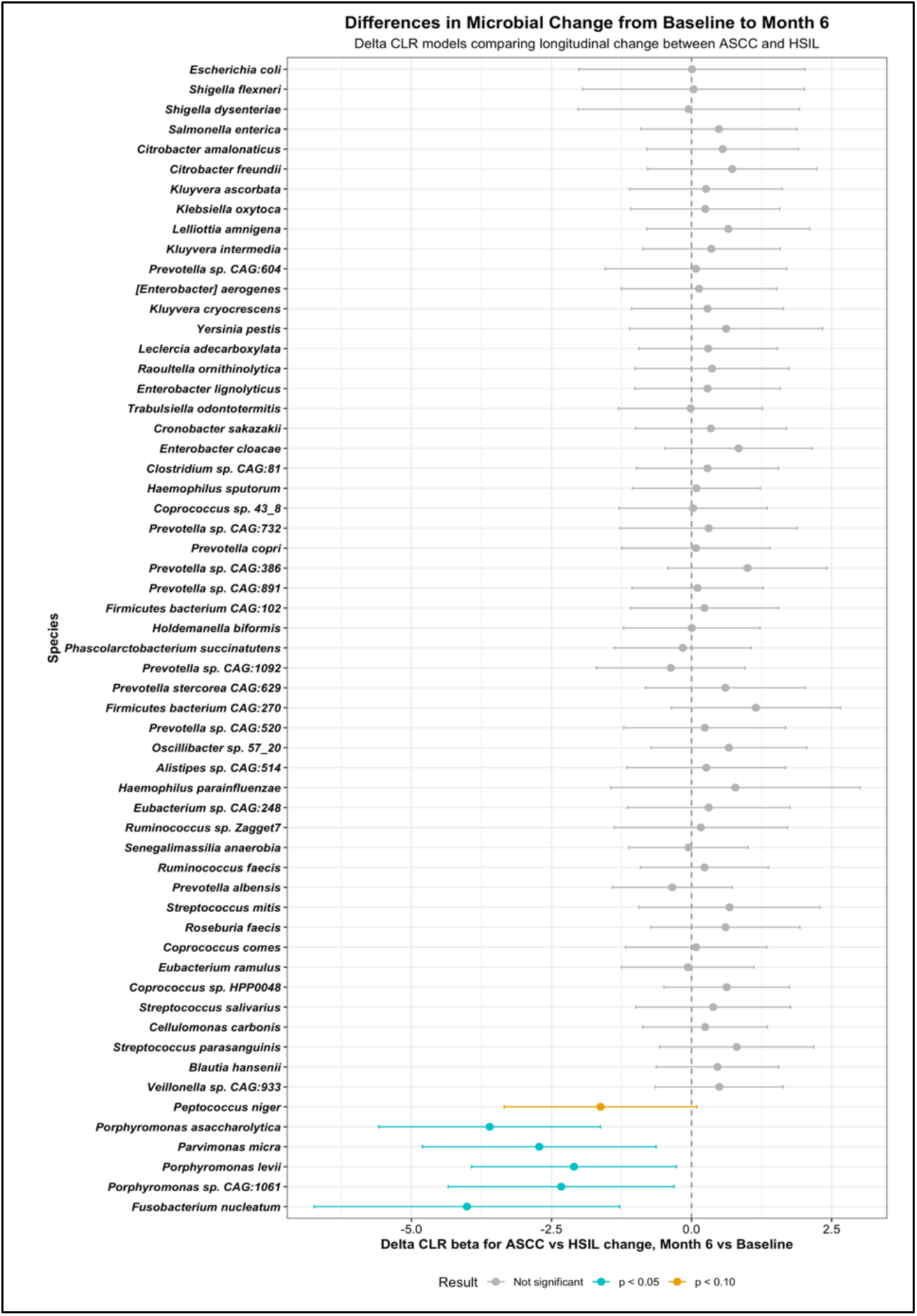
ASCC-associated anaerobic species decrease preferentially from baseline to Month 6. Forest plot showing differences between ASCC and HSIL in the change in centered log-ratio (CLR) abundance from baseline to Month 6 among validated baseline candidate species. Delta CLR was calculated as Month 6 CLR abundance minus baseline CLR abundance for each participant. Points represent regression coefficients comparing temporal change in ASCC with that in HSIL, and horizontal lines represent 95% confidence intervals. Negative coefficients indicate a greater decrease from baseline to Month 6 in ASCC relative to HSIL. Five ASCC-enriched species, *Fusobacterium nucleatum*, *Porphyromonas asaccharolytica*, *Parvimonas micra*, *Porphyromonas* sp. CAG:1061, and *Porphyromonas levii*, demonstrated significantly greater decreases in ASCC. Colors indicate associations with *p* < 0.05, *p* < 0.10, or nonsignificant associations, as indicated in the figure. The vertical dashed line at β = 0 indicates no difference in temporal change between groups.

### ASCC-Only Baseline-to-Month 2 Changes

To assess whether these decreases were already emerging during the earlier treatment interval, exploratory paired ASCC-only baseline-to-Month 2 analyses were performed among 14 patients. Among the 58 validated baseline species, 26 decreased and 32 increased from Baseline to Month 2. The strongest early decreases were again observed among ASCC-associated anaerobic taxa. Parvimonas micra decreased significantly from baseline to Month 2 (β = −2.91, 95% CI −4.86 to −0.96, p = 0.0067), as did Porphyromonas levii (β = −1.78, 95% CI −3.50 to −0.07, p = 0.0425). Additional ASCC-enriched taxa showed similar directional decreases, including Fusobacterium nucleatum (β = −2.86, 95% CI −5.73 to 0.00, p = 0.0503), Porphyromonas asaccharolytica (β = −1.92, 95% CI −3.94 to 0.10, p = 0.0607), and Peptococcus niger (β = −1.08, 95% CI −2.29 to 0.13, p = 0.0757). Although Month 2 samples were available only for ASCC participants, these paired analyses support the same directional pattern observed at Month 6, with early reductions in several baseline ASCC-enriched anaerobes following treatment initiation.

### Community-Level Functional Associations

Functional analyses evaluated 152 MetaCyc pathways in relation to the six persistent ASCC-enriched species. Twenty-six pathways were positively associated with at least one of these species and were independently enriched in ASCC after covariate adjustment and FDR correction in both models (Figure 6). Of these, 22 pathways were associated with at least two species, 15 with at least three species, and nine with all six species. Shared pathways were primarily related to amino acid metabolism, carbohydrate and energy metabolism, and anaerobic degradation, including L-histidine degradation, L-glutamate degradation, L-aspartate degradation, lysine and homoserine biosynthesis, and pyruvate fermentation to butanoate. Collectively, these findings identified a shared community-level functional profile associated with persistent ASCC-enriched taxa, characterized primarily by amino acid metabolism and anaerobic metabolic pathways.

**Figure 6.**
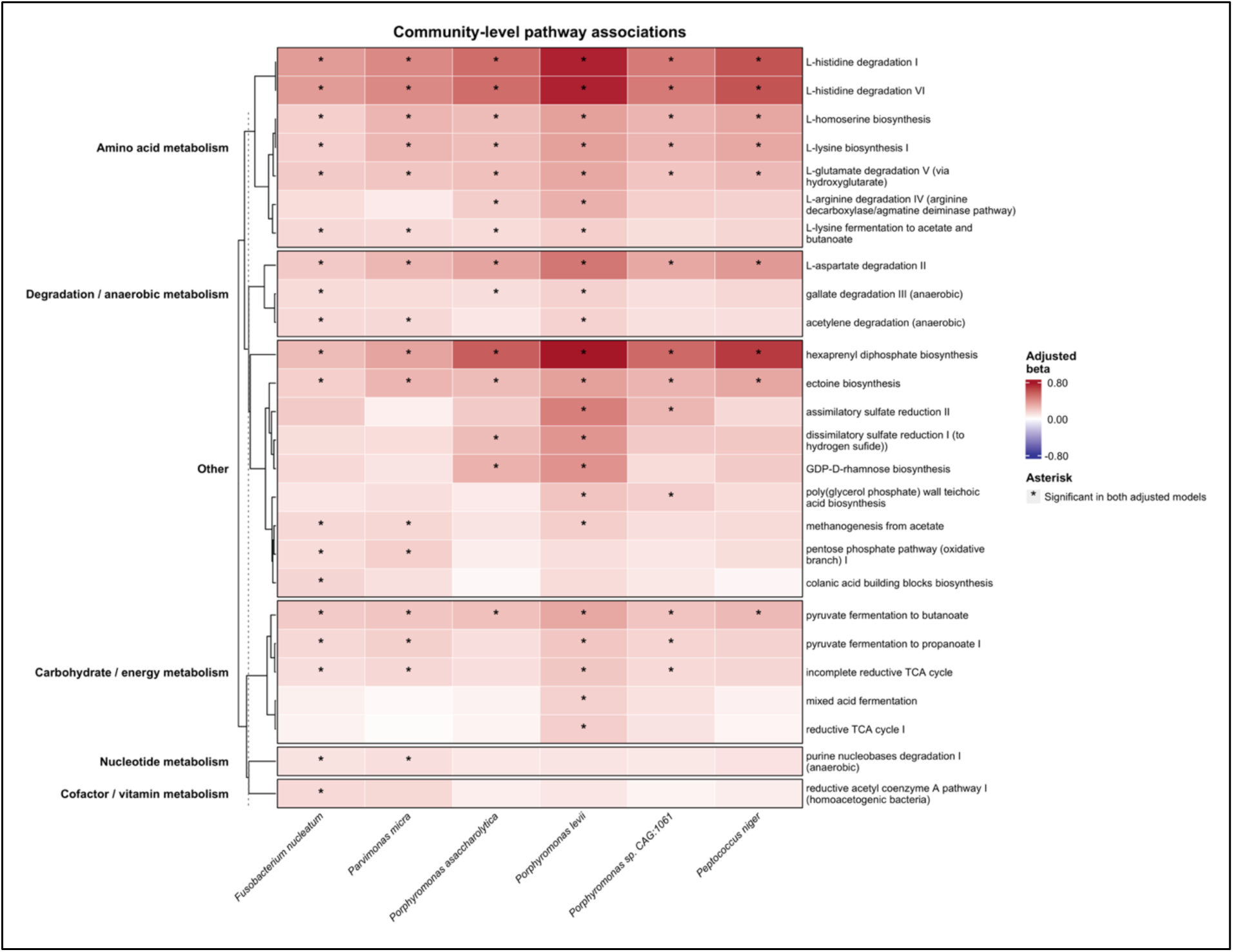
Persistent ASCC-enriched species share a community-level functional profile dominated by amino acid and anaerobic metabolism. Heatmap showing community-level MetaCyc pathway associations with the six species that remained enriched in anal squamous cell carcinoma (ASCC) across longitudinal sampling: *Fusobacterium nucleatum*, *Parvimonas micra*, *Porphyromonas asaccharolytica*, *Porphyromonas levii*, *Porphyromonas* sp. CAG:1061, and *Peptococcus niger*. Among 152 pathways evaluated, 26 were positively associated with at least one persistent ASCC-enriched species and were independently enriched in ASCC after adjustment for age, sex, tobacco use, HIV status, and HPV status and FDR correction. Rows represent these 26 MetaCyc pathways and columns represent the six persistent ASCC-enriched species. Cell color represents the adjusted regression coefficient for the association between species CLR abundance and pathway CLR abundance. Asterisks identify species-pathway associations for which the species-pathway association and independent ASCC-versus-HSIL pathway comparison both met an FDR-adjusted q value < 0.05. Twenty-two pathways were associated with at least two species, 15 with at least three species, and nine with all six species. Pathways are grouped by functional category and were predominantly related to amino acid metabolism, anaerobic degradation, and carbohydrate and energy metabolism. Because pathway abundances were measured at the community level, these associations do not establish species-specific functional contributions.

## Discussion

In this prospective study, we found that the anal microbiome differed between patients with HSIL and those with non-metastatic ASCC. ASCC was associated with lower observed species richness, although Shannon and inverse Simpson diversity were similar between groups. Community composition also differed across Bray-Curtis, Jaccard, and Aitchison metrics after adjustment for age, sex, tobacco use, HIV status, and HPV status. At the species level, ASCC was characterized by enrichment of a consistent anaerobic cluster comprising Fusobacterium nucleatum, Parvimonas micra, Porphyromonas asaccharolytica, Porphyromonas levii, Porphyromonas sp. CAG:1061, and Peptococcus niger. These taxa were identified using complementary compositional approaches, appeared among the most important features in the exploratory random forest model, and remained associated with ASCC across longitudinal sampling. Five also decreased more from baseline to Month 6 in ASCC than in HSIL, with early decreases evident by Month 2. Beyond these taxonomic differences, functional analyses linked the persistent ASCC-enriched taxa to a shared community-level profile dominated by amino acid metabolism and anaerobic metabolic pathways.

Our findings are consistent with prior studies showing differences in the anorectal microbiome across HPV-related anal disease.^14–16^ Elnaggar et al. reported differences in microbial composition among individuals with HPV infection, anal dysplasia, and anal cancer, with enrichment of potentially pathogenic and inflammatory taxa in cancer. However, the small number of cancer cases and use of 16S rRNA sequencing limited species-level resolution.^14^ More recently, metatranscriptomic profiling demonstrated separation of microbial communities across low-grade lesions, HSIL, and ASCC and identified enrichment of F. nucleatum, Bacteroides fragilis, and Campylobacter ureolyticus in invasive disease.^16^ That study also identified microbial proteins and virulence-associated factors, including the F. nucleatum adhesin FadA, suggesting that some ASCC-associated organisms may be active within the tumor microenvironment rather than representing incidental surface colonization.^16^

Our alpha-diversity findings differ from some prior reports. We observed lower species richness in ASCC, whereas a recent metatranscriptomic analysis reported greater richness and diversity in ASCC than in HSIL.^16^ Other studies found similar alpha diversity between dysplasia and cancer despite differences in individual taxa and overall community composition.^14^ These differences may be related to the sampling site, specimen type, sequencing platform, cohort characteristics, preprocessing methods, treatment exposure, or taxonomic filtering. Metatranscriptomic sequencing evaluates transcriptionally active organisms in tissue,^24^ whereas shotgun metagenomic sequencing of anal canal swabs detects microbial DNA from viable, inactive, and transient organisms. Notably, the anal canal microbiome is compositionally distinct from stool, supporting site-specific sampling in this setting.^15^ Alpha diversity may also be less informative than community-level and species-level analyses because it reduces complex ecological differences to a single summary measure. In our cohort, disease group explained 10.0% to 17.2% of community variation across adjusted beta-diversity models. The absence of dispersion differences for Bray-Curtis and Aitchison distances supports that these findings reflect differences in community composition rather than greater variability within one group.

The clearest ASCC-associated finding was enrichment of a small group of anaerobic organisms, several of which have also been reported in colorectal and other gastrointestinal cancers. ^25–27^ F. nucleatum and P. micra are among the most consistently enriched bacterial species in colorectal cancer, and a systematic review of shotgun metagenomic studies identified them as the two most reproducibly reported CRC-associated taxa.^27^ Similarly, multimicrobial panels incorporating both organisms have shown promising accuracy for distinguishing patients with colorectal cancer from non-CRC controls.^28–30^ Porphyromonas asaccharolytica has also been identified as a cancer-associated species in studies using quantitative microbial profiling and adjustment for relevant covariates.^31^ The concurrent enrichment of Fusobacterium, Parvimonas, Porphyromonas, and Peptococcus may therefore be more biologically meaningful than the presence of any single organism, suggesting a shared anaerobic microbial community rather than isolated colonizers.^30,32^

The longitudinal analysis further supported the reproducibility of this ASCC-associated signature. All six ASCC-enriched anaerobic species identified by both ANCOM-BC2 and CLR regression remained associated with disease group across baseline, Month 6, and Month 12, whereas most of the broader baseline HSIL-associated signature did not remain significant over repeated sampling. Five of the six principal ASCC-enriched species also decreased significantly more from baseline to Month 6 in ASCC than in HSIL. In the ASCC-only Month 2 analysis, P. micra and P. levii decreased significantly, while F. nucleatum, P. asaccharolytica, and P. niger showed similar directional changes. Thus, the taxa most strongly enriched before treatment were also those that decreased most clearly after treatment began. An analogous reduction in cancer-associated taxa after curative treatment has been reported in colorectal cancer.^25,27^ Although this pattern is consistent with a relationship between the microbial signature and the tumor-associated environment, the observed changes cannot be attributed specifically to treatment effects or tumor response, as chemoradiotherapy, antibiotic exposure, dietary changes, bowel dysfunction, and treatment-related mucosal inflammation may also alter the anal microbiome. Prior longitudinal studies have similarly documented substantial shifts in the anorectal microbiome during and after chemoradiation for ASCC.^33^

The exploratory random forest model distinguished ASCC from HSIL with an AUC of 0.844, and several of the highest-ranked features overlapped with the ANCOM-BC2 and CLR-validated signature. This finding supports the potential discriminatory value of the identified taxa, although the classifier remains hypothesis-generating. The ASCC cohort was small, leave-one-out cross-validation does not replace external validation, and model performance may decline across different populations, sequencing platforms, and sampling methods. The more clinically useful application may not be distinguishing established HSIL from established ASCC, which is determined histologically, but rather identifying patients with HSIL who are at greatest risk of persistence, recurrence, or progression. This approach is supported by prior work demonstrating that microbial features improve the specificity of anal cytology-based screening.^34–36^ Future studies should evaluate whether this microbial signature precedes progression and whether its integration with HPV genotype, immune markers, and lesion-specific molecular features improves risk stratification.^16^

Community-level functional analysis complements the taxonomic signature by identifying metabolic pathways associated with the persistent ASCC-enriched species. These findings also raise important mechanistic questions. The co-occurrence of F. nucleatum, P. micra, multiple Porphyromonas species, and P. niger suggests that ASCC may support a coordinated anaerobic community rather than independent expansion of individual taxa.^30,32^ It remains unknown whether these organisms are present within tumor tissue, limited to the mucosal surface, or organized within biofilms. Studies using paired anal swabs, stool, tumor, and adjacent normal tissue could help distinguish a local tumor-associated community from broader gastrointestinal microbial patterns.^15^ Metatranscriptomic, metabolomic, and spatial approaches could further determine whether these organisms are active and whether their presence is associated with hypoxia, inflammation, immune-cell composition, or treatment response.^16,24^ Functional studies will ultimately be needed to determine whether the identified organisms affect HPV persistence, malignant transformation, antitumor immunity, or radiosensitivity.^37^

This study has several limitations. Although the overall cohort was relatively large for an anal microbiome study, only 19 patients with ASCC had evaluable baseline samples, resulting in group imbalance and limited precision for cancer-specific estimates. Patients with ASCC were older, while HIV positivity was more common in the HSIL group. We adjusted for age, sex, tobacco use, HIV status, and HPV status, but residual confounding remains possible. Other factors, including antibiotic exposure, sexual practices, bowel preparation, diet, body mass index, stool consistency, local inflammation, antiretroviral therapy, and other medications, may also affect microbial composition and were not included in the reported models. Host factors such as stool consistency and transit time, intestinal inflammation, and body mass index have been shown to be important determinants of microbial composition and should be considered in future validation studies.^31^

The analysis was also based on relative abundance and therefore cannot determine whether an enriched organism increased in absolute abundance or appeared enriched because other taxa decreased. Quantitative microbial profiling, internal spike-in standards, or targeted quantitative PCR would strengthen future studies. Longitudinal analyses were limited to available participants and specimens and may be affected by informative missingness. Month 2 samples were available only for ASCC, and the small paired cohort (n = 14) limited power to detect early changes. In addition, this observational study cannot determine whether the identified microbial community contributes to malignant progression or develops because of invasive disease.

In conclusion, ASCC was associated with a distinct anal microbial community characterized by lower observed richness and enrichment of a reproducible anaerobic cluster containing F. nucleatum, P. micra, P. asaccharolytica, P. levii, Porphyromonas sp. CAG:1061, and P. niger. This signature remained associated with ASCC across repeated sampling but decreased after treatment initiation. These findings support a close relationship between the local microbial environment and invasive anal cancer. Future studies should build on these findings to evaluate the identified taxa as markers of HSIL progression, tumor burden, or treatment response and to define their potential functional role in HPV-driven carcinogenesis.

## Ethics Approval and Consent to Participate

This study was conducted in accordance with institutional review board approval at Montefiore Medical Center. Written informed consent was obtained from all participants prior to specimen collection.

## Data Availability

Data supporting the findings of this study are available from the corresponding author upon reasonable request.

---

**Supplementary Figure 1.**
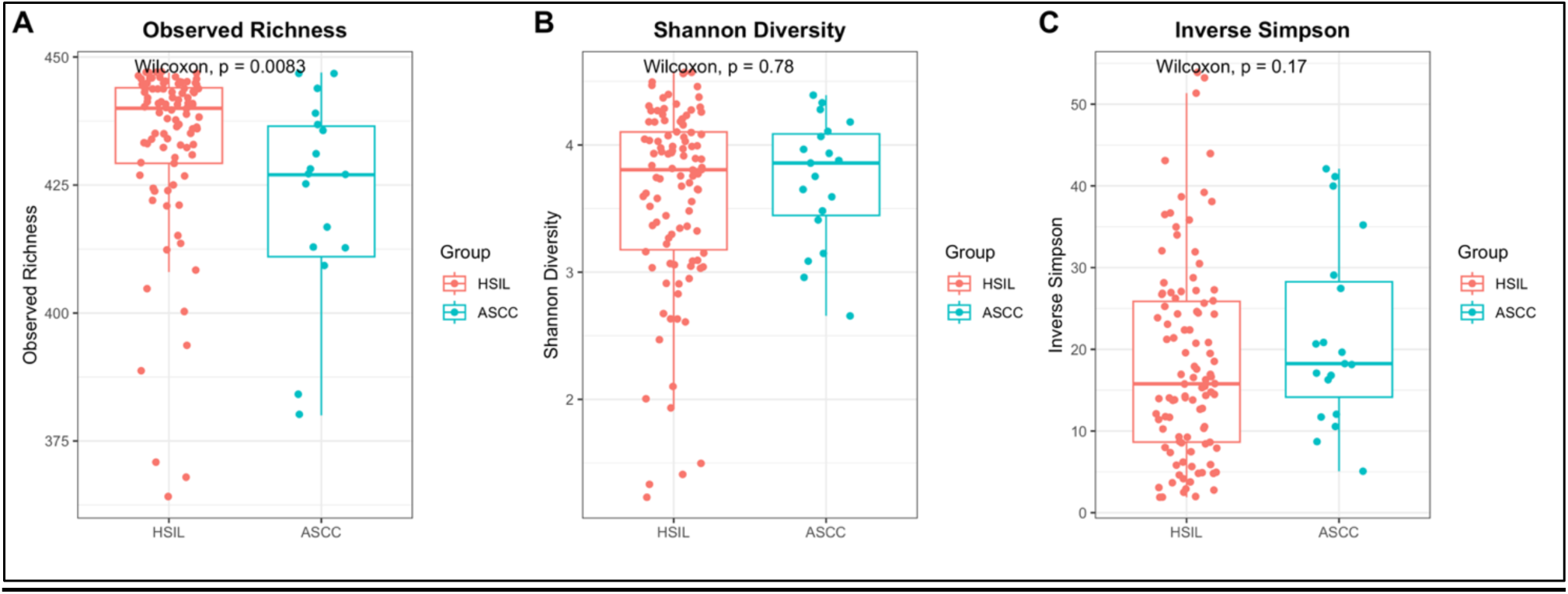
Species-level alpha diversity in HSIL and ASCC. Species-level alpha diversity in baseline anal canal microbiome samples from patients with HSIL (n = 102) and ASCC (n = 19). (A) Observed species richness, (B) Shannon diversity, and (C) inverse Simpson diversity were calculated from core-filtered raw count data. Boxplots show the median and interquartile range, with individual samples overlaid. Between-group differences were evaluated using Wilcoxon rank-sum tests. Observed richness was significantly lower in ASCC, whereas Shannon and inverse Simpson diversity did not differ significantly between groups.

**Supplementary Figure 2.**
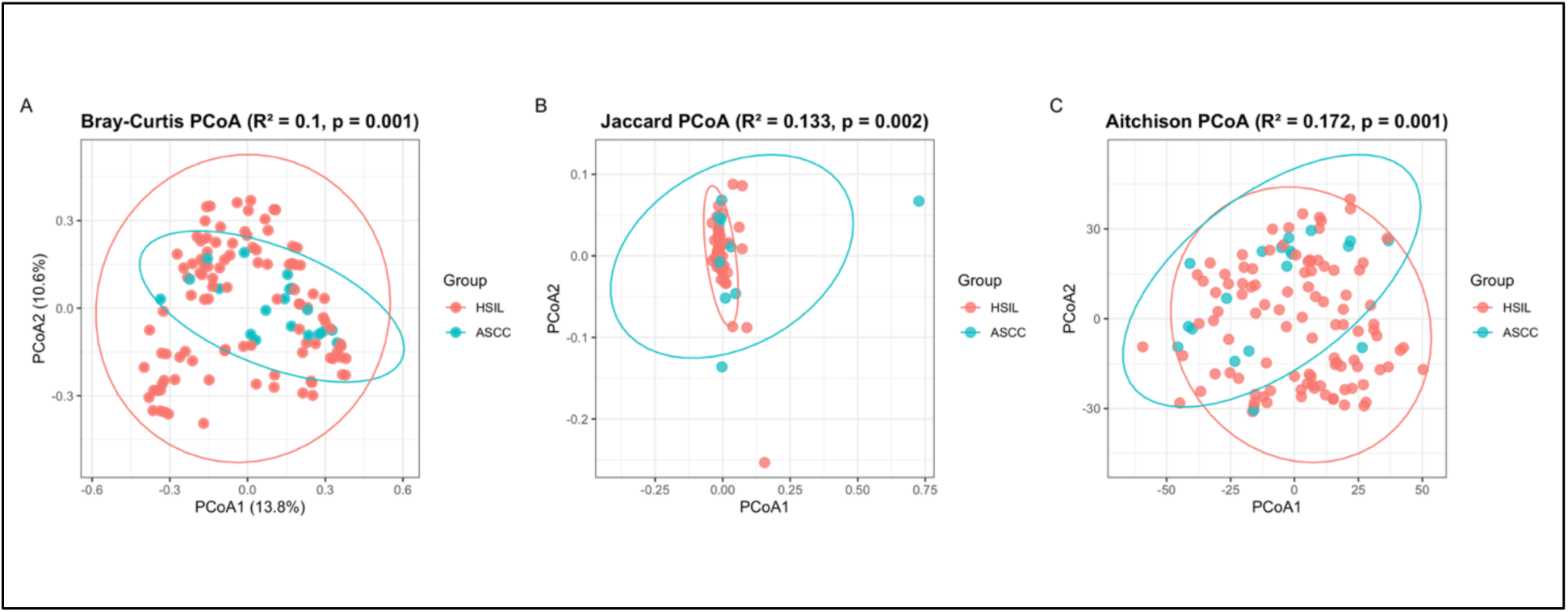
Beta-diversity analyses demonstrate differences in microbial community composition between HSIL and ASCC. Principal coordinates analyses of baseline anal microbial community composition comparing HSIL and ASCC using (A) Bray-Curtis dissimilarity, (B) Jaccard distance, and (C) Aitchison distance derived from centered log-ratio-transformed abundances. Each point represents an individual baseline sample, and ellipses depict the distribution of samples within each disease group. Covariate-adjusted PERMANOVA models with 999 permutations demonstrated significant separation by disease group for Bray-Curtis (R² = 0.100, p = 0.001), Jaccard (R² = 0.133, p = 0.002), and Aitchison (R² = 0.172, p = 0.001) distances. Models were adjusted for age, sex, tobacco use, HIV status, and HPV status.

**Supplementary Figure 3.**
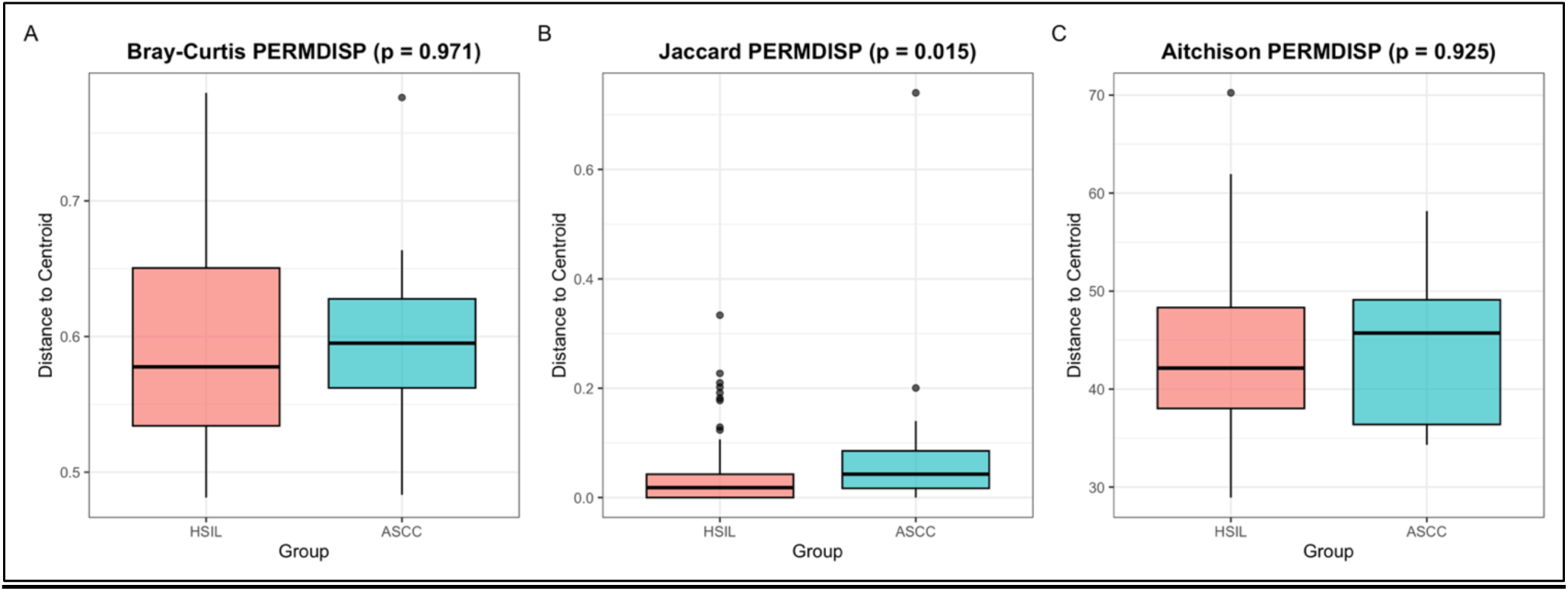
Multivariate dispersion across beta-diversity metrics in HSIL and ASCC. Permutational analysis of multivariate dispersions (PERMDISP) comparing within-group dispersion between HSIL and ASCC using (A) Bray–Curtis dissimilarity, (B) Jaccard distance, and (C) Aitchison distance. Boxplots show the distribution of distances from individual samples to the corresponding group centroid. No significant differences in dispersion were observed for Bray–Curtis (*p* = 0.971) or Aitchison (*p* = 0.925) distances, supporting the interpretation that disease-associated differences using these metrics reflected differences in microbial community structure rather than dispersion alone. Jaccard dispersion differed significantly between groups (*p* = 0.015), and disease-associated Jaccard results were therefore interpreted more cautiously.

**Supplementary Figure 4.**
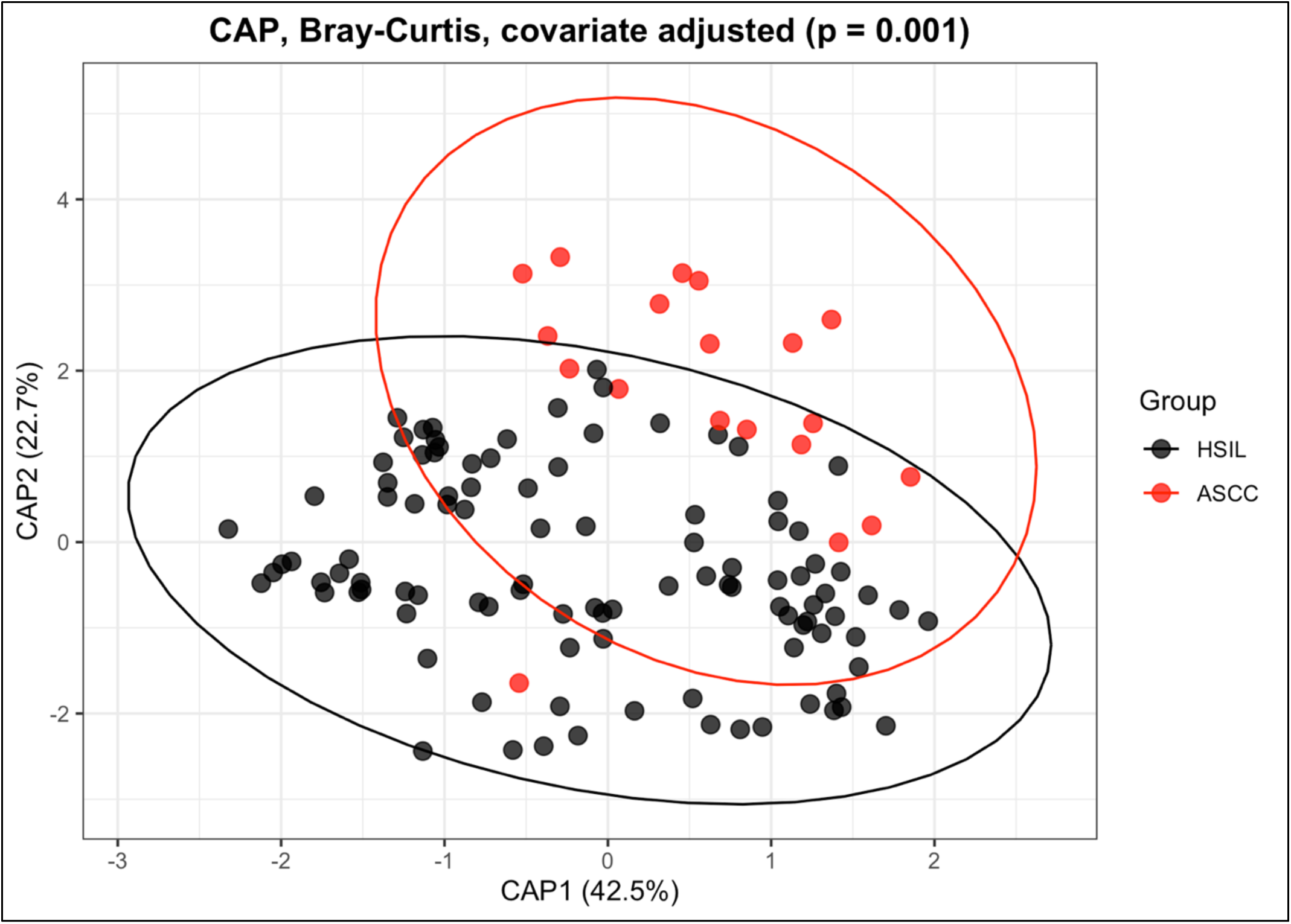
Covariate-adjusted constrained ordination demonstrates disease-associated microbial community separation. Constrained analysis of principal coordinates (CAP) using Bray–Curtis dissimilarity to evaluate differences in baseline microbial community composition between patients with HSIL and ASCC. The analysis was adjusted for age, sex, tobacco use, HIV status, and HPV status. Each point represents an individual baseline sample, and ellipses depict the distribution of samples within each disease group. Permutation-based testing demonstrated significant disease-associated community separation (*p* = 0.001).

**Supplementary Figure 5.**
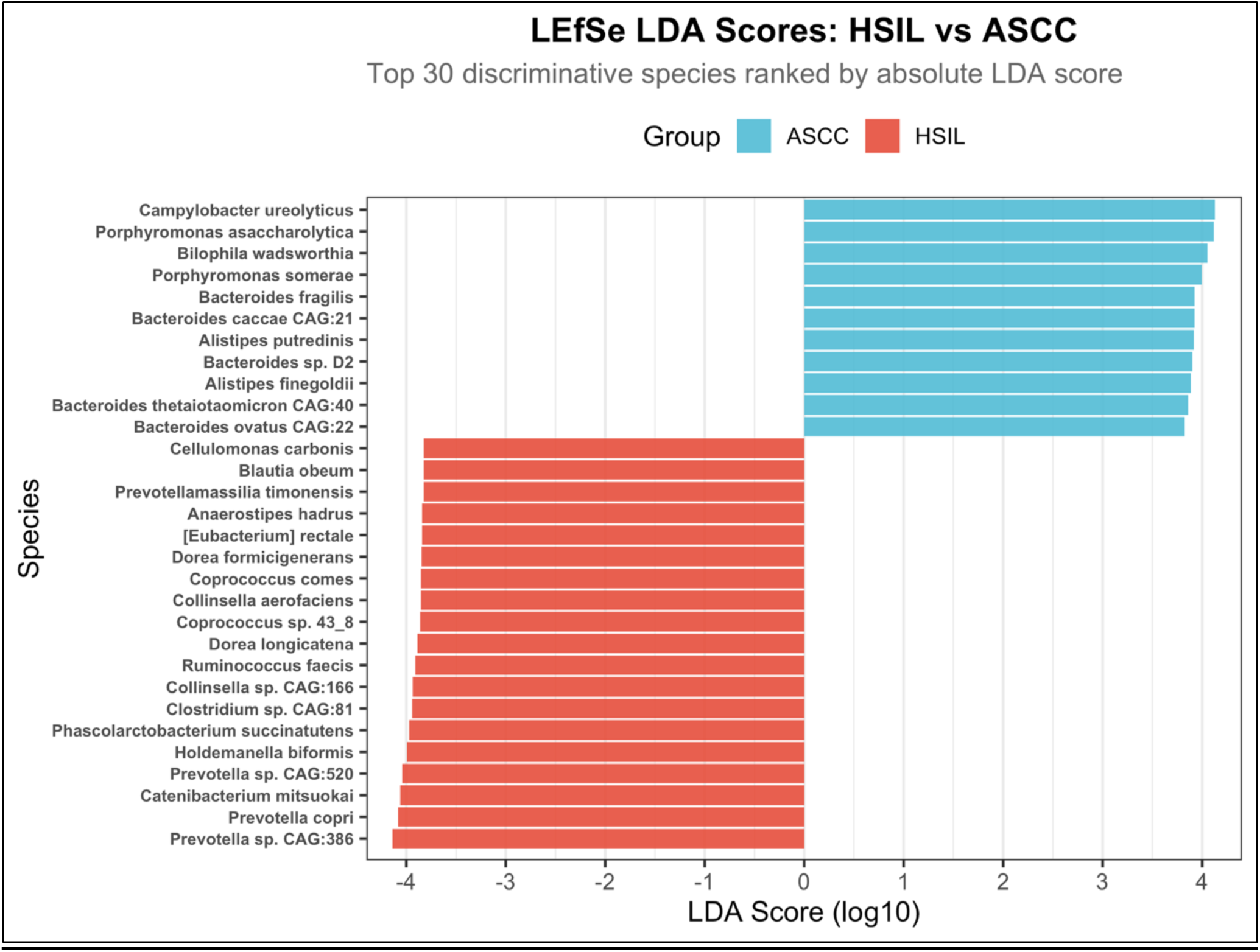
Exploratory LEfSe analysis identifies species discriminating HSIL from ASCC. Linear discriminant analysis effect size (LEfSe) analysis of baseline species-level microbial profiles from patients with HSIL and ASCC. The 30 species with the largest absolute linear discriminant analysis (LDA) scores are shown. Positive LDA scores indicate enrichment in ASCC, whereas negative LDA scores indicate enrichment in HSIL. LEfSe was performed as a complementary exploratory analysis and was not used for primary candidate-species selection.

**Supplementary Figure 6.**
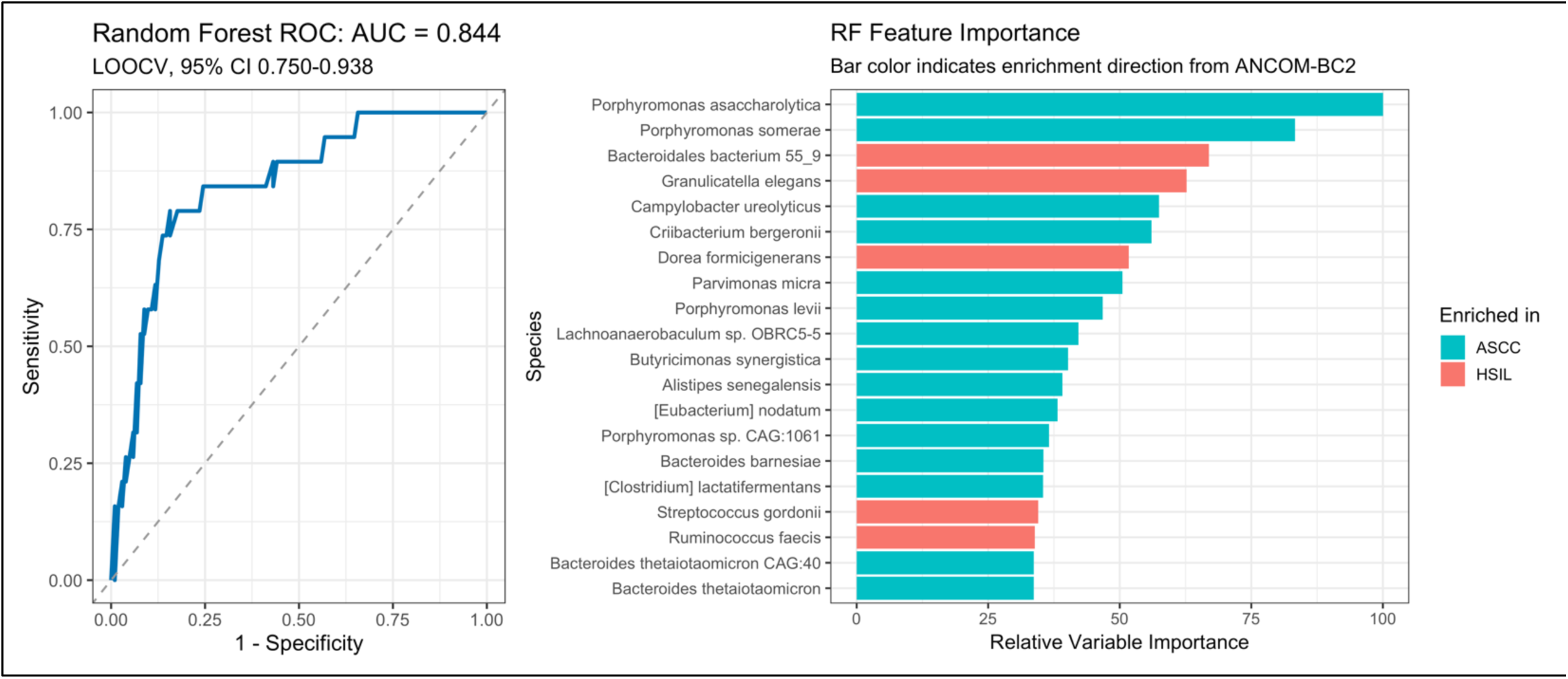
Exploratory random forest classification distinguishes HSIL from ASCC using baseline microbial features. (A) Receiver operating characteristic (ROC) curve for a random forest classifier trained using baseline species-level microbial abundance profiles to distinguish HSIL from ASCC. Model performance was evaluated using leave-one-out cross-validation (LOOCV), yielding an area under the ROC curve (AUC) of 0.844 (95% CI, 0.750–0.938). The diagonal dashed line represents discrimination expected by chance. (B) Relative variable importance of the highest-ranked microbial features contributing to random forest classification. Higher values indicate a greater contribution to classification. Bar color indicates the direction of enrichment identified by ANCOM-BC2, with taxa enriched in ASCC and HSIL shown separately. Random forest analysis was considered exploratory and was not used for primary candidate-species selection.

**Supplementary Table S1.**
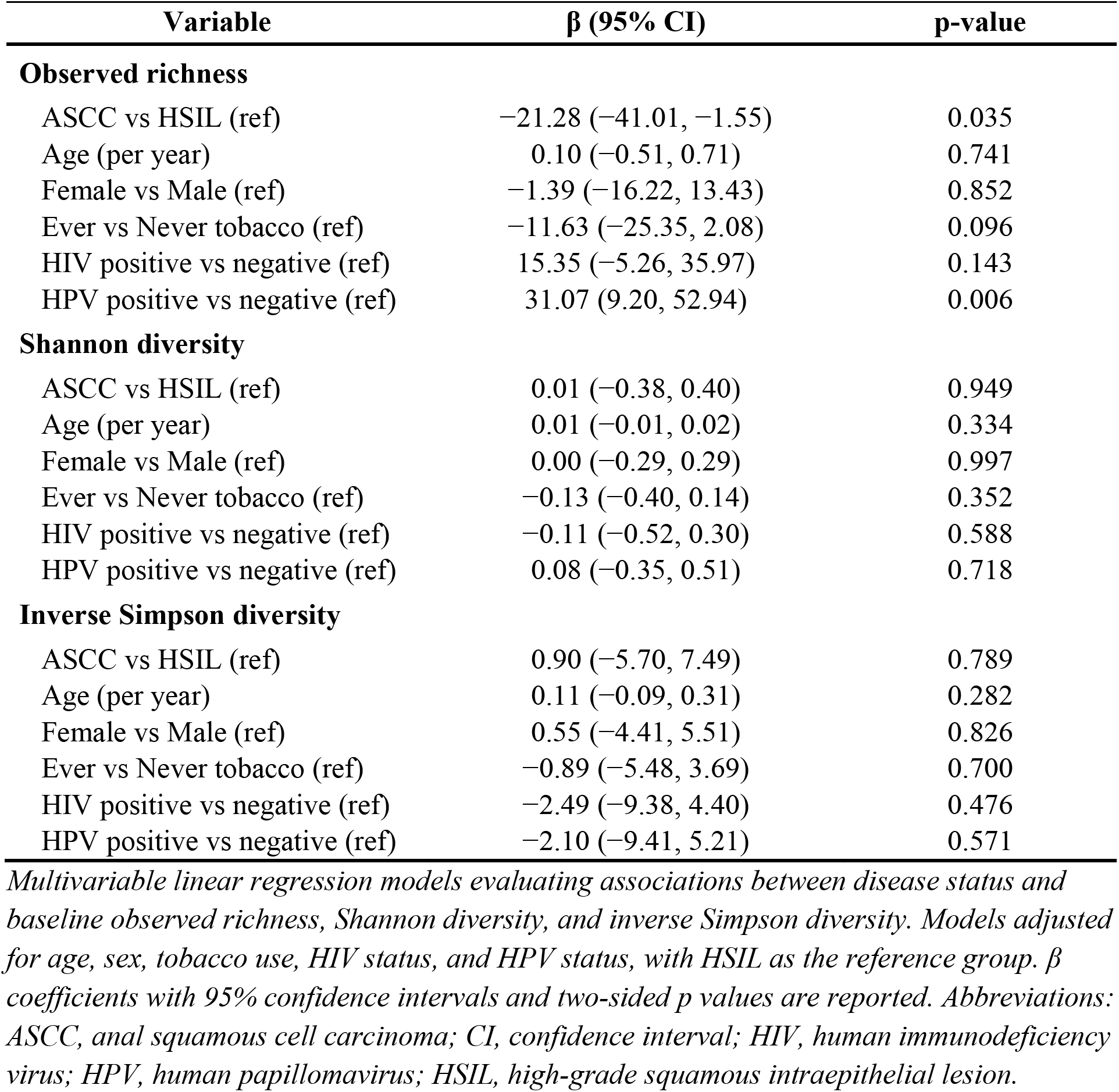
Multivariable linear regression of alpha diversity metrics at baseline.

| Variable | $\beta$ (95% CI) | p-value |
| --- | --- | --- |
| **Observed richness** |  |  |
| ASCC vs HSIL (ref) | -21.28 (-41.01, -1.55) | 0.035 |
| Age (per year) | 0.10 (-0.51, 0.71) | 0.741 |
| Female vs Male (ref) | -1.39 (-16.22, 13.43) | 0.852 |
| Ever vs Never tobacco (ref) | -11.63 (-25.35, 2.08) | 0.096 |
| HIV positive vs negative (ref) | 15.35 (-5.26, 35.97) | 0.143 |
| HPV positive vs negative (ref) | 31.07 (9.20, 52.94) | 0.006 |
| **Shannon diversity** |  |  |
| ASCC vs HSIL (ref) | 0.01 (-0.38, 0.40) | 0.949 |
| Age (per year) | 0.01 (-0.01, 0.02) | 0.334 |
| Female vs Male (ref) | 0.00 (-0.29, 0.29) | 0.997 |
| Ever vs Never tobacco (ref) | -0.13 (-0.40, 0.14) | 0.352 |
| HIV positive vs negative (ref) | -0.11 (-0.52, 0.30) | 0.588 |
| HPV positive vs negative (ref) | 0.08 (-0.35, 0.51) | 0.718 |
| **Inverse Simpson diversity** |  |  |
| ASCC vs HSIL (ref) | 0.90 (-5.70, 7.49) | 0.789 |
| Age (per year) | 0.11 (-0.09, 0.31) | 0.282 |
| Female vs Male (ref) | 0.55 (-4.41, 5.51) | 0.826 |
| Ever vs Never tobacco (ref) | -0.89 (-5.48, 3.69) | 0.700 |
| HIV positive vs negative (ref) | -2.49 (-9.38, 4.40) | 0.476 |
| HPV positive vs negative (ref) | -2.10 (-9.41, 5.21) | 0.571 |

*Multivariable linear regression models evaluating associations between disease status and baseline observed richness, Shannon diversity, and inverse Simpson diversity. Models adjusted for age, sex, tobacco use, HIV status, and HPV status, with HSIL as the reference group. $\beta$ coefficients with 95% confidence intervals and two-sided p values are reported. Abbreviations: ASCC, anal squamous cell carcinoma; CI, confidence interval; HIV, human immunodeficiency virus; HPV, human papillomavirus; HSIL, high-grade squamous intraepithelial lesion.*

**Supplementary Table S2.** All 60 FDR-significant species identified by ANCOM-BC2.

| Species | Log fold change | FDR q-value | Enriched group |
| --- | --- | --- | --- |
| *Escherichia coli* | -3.59 | 0.00685 | HSIL |
| *Prevotella* sp. CAG:604 | -3.55 | 0.000209 | HSIL |
| *Fusobacterium nucleatum* | 3.39 | 0.00685 | ASCC |
| *Porphyromonas asaccharolytica* | 3.25 | 0.0168 | ASCC |
| *Porphyromonas* sp. CAG:1061 | 3.23 | 0.0116 | ASCC |
| *Shigella flexneri* | -3.19 | 0.00685 | HSIL |
| *Shigella dysenteriae* | -3.17 | 0.00754 | HSIL |
| *Parvimonas micra* | 2.99 | 0.00946 | ASCC |
| *Citrobacter amalonaticus* | -2.95 | 0.00554 | HSIL |
| *Acidaminococcus intestini* CAG:325 | 2.94 | 0.00485 | ASCC |
| *Salmonella enterica* | -2.87 | 0.00554 | HSIL |
| *Prevotella* sp. CAG:386 | -2.85 | 0.00685 | HSIL |
| *Porphyromonas levii* | 2.82 | 0.00685 | ASCC |
| *Prevotella* sp. CAG:732 | -2.78 | 0.00554 | HSIL |
| *Lelliottia amnigena* | -2.73 | 0.00554 | HSIL |
| *Yersinia pestis* | -2.68 | 0.0145 | HSIL |
| *Trabulsiella odontotermitis* | -2.54 | 0.00554 | HSIL |
| *Prevotella stercorea* CAG:629 | -2.52 | 0.00554 | HSIL |
| [*Enterobacter*] *aerogenes* | -2.45 | 0.00685 | HSIL |
| *Citrobacter freundii* | -2.40 | 0.00902 | HSIL |
| *Kluyvera ascorbata* | -2.37 | 0.00554 | HSIL |
| *Enterobacter lignolyticus* | -2.35 | 0.00685 | HSIL |
| *Prevotella* sp. CAG:520 | -2.34 | 0.00554 | HSIL |
| *Alistipes* sp. CAG:514 | -2.33 | 0.00554 | HSIL |
| *Haemophilus parainfluenzae* | -2.27 | 0.0163 | HSIL |
| *Prevotella copri* | -2.25 | 0.0238 | HSIL |
| *Kluyvera cryocrescens* | -2.24 | 0.0102 | HSIL |
| *Firmicutes bacterium* CAG:270 | -2.20 | 0.0210 | HSIL |
| *Enterobacter cloacae* | -2.19 | 0.0104 | HSIL |
| *Raoultella ornithinolytica* | -2.16 | 0.0104 | HSIL |
| *Firmicutes bacterium* CAG:102 | -2.14 | 0.0482 | HSIL |
| *Clostridium* sp. CAG:81 | -2.11 | 0.00554 | HSIL |
| *Prevotella albensis* | -2.09 | 0.00685 | HSIL |
| *Ruminococcus* sp. Zagget7 | -2.05 | 0.0210 | HSIL |
| *Kluyvera intermedia* | -2.04 | 0.0187 | HSIL |
| *Oscillibacter* sp. 57_20 | -2.04 | 0.0479 | HSIL |
| *Haemophilus sputorum* | -2.02 | 0.0112 | HSIL |
| *Leclercia adecarboxylata* | -2.00 | 0.0143 | HSIL |
| *Klebsiella oxytoca* | -1.98 | 0.0210 | HSIL |
| *Prevotella* sp. CAG:891 | -1.97 | 0.00554 | HSIL |
| *Coprococcus* sp. 43_8 | -1.96 | 0.0116 | HSIL |
| Veillonella sp. CAG:933 | -1.92 | 0.0152 | HSIL |
| Streptococcus salivarius | -1.91 | 0.0116 | HSIL |
| Roseburia faecis | -1.87 | 0.0452 | HSIL |
| Prevotella sp. CAG:1092 | -1.86 | 0.0284 | HSIL |
| Phascolarctobacterium succinatutens | -1.86 | 0.0327 | HSIL |
| Cronobacter sakazakii | -1.85 | 0.0241 | HSIL |
| Streptococcus mitis | -1.81 | 0.0325 | HSIL |
| Peptococcus niger | 1.80 | 0.0375 | ASCC |
| Ruminococcus faecis | -1.76 | 0.0325 | HSIL |
| Streptococcus parasanguinis | -1.74 | 0.0325 | HSIL |
| Eubacterium sp. CAG:248 | -1.71 | 0.0323 | HSIL |
| Holdemanella biformis | -1.71 | 0.0311 | HSIL |
| Coprococcus comes | -1.71 | 0.0284 | HSIL |
| Allisonella histaminiformans | -1.66 | 0.0299 | HSIL |
| Coprococcus sp. HPP0048 | -1.64 | 0.0287 | HSIL |
| Senegalimassilia anaerobia | -1.63 | 0.0299 | HSIL |
| Cellulomonas carbonis | -1.53 | 0.0408 | HSIL |
| Blautia hansenii | -1.53 | 0.0211 | HSIL |
| Eubacterium ramulus | -1.38 | 0.0482 | HSIL |

*Species meeting FDR-adjusted significance in differential abundance analysis comparing ASCC and HSIL at baseline. Log fold change reflects relative abundance in ASCC versus HSIL; positive values indicate enrichment in ASCC and negative values indicate enrichment in HSIL. FDR-adjusted q values are reported. Abbreviations: ANCOM-BC2, Analysis of Compositions of Microbiomes with Bias Correction 2; ASCC, anal squamous cell carcinoma; FDR, false discovery rate; HSIL, high-grade squamous intraepithelial lesion.*

